# An Objective Structural and Functional Reference Standard for Diagnostic Studies in Glaucoma

**DOI:** 10.1101/2020.04.10.20057836

**Authors:** Eduardo B. Mariottoni, Alessandro A. Jammal, Samuel I. Berchuck, Ivan M. Tavares, Felipe A. Medeiros

## Abstract

**Purpose:** To propose a reference standard for the definition of glaucomatous optic neuropathy (GON) consisting of objective parameters from spectral-domain optical coherence tomography (SDOCT) and standard automated perimetry (SAP), and to apply it to the development and evaluation of a deep learning (DL) algorithm to detect glaucomatous damage on fundus photographs.

**Design:** Retrospective, cross-sectional study.

**Methods:** Data were extracted from the Duke Glaucoma Registry and included 2,927 eyes of 2,025 participants with fundus photos, SDOCT and SAP acquired within six months. Eyes were classified as GON versus normal based on a combination of objective SDOCT and SAP criteria. A DL convolutional neural network was trained to predict the probability of GON from fundus photos. The algorithm was tested on an independent sample with performance assessed by sensitivity, specificity, area under the receiver operating characteristic curve (AUC), and likelihood ratios (LR).

**Results:** The test sample included 585 eyes of 405 participants. The median DL probability of glaucoma in eyes with GON was 99.8% versus 0.03% for normal eyes (P < 0.001), with an AUC of 0.92 and sensitivity of 77% at 95% specificity. LRs indicated that the DL algorithm provided large changes in the post-test probability of disease for the majority of eyes.

**Conclusions:** The DL algorithm had high performance to discriminate eyes with GON from normal. The newly proposed objective definition of GON used as reference standard may increase the comparability of diagnostic studies of glaucoma across devices and populations, helping to improve the development and assessment of tests in clinical practice.

## 1 Introduction

Glaucoma is a progressive optic neuropathy in which characteristic changes in the optic disc and retinal nerve fiber layer (RNFL) are usually accompanied by associated visual field defects.[1] The disease is the leading cause of irreversible blindness, affecting over 70 million people worldwide.[2] Nonetheless, despite recent advances in functional and structural assessment, diagnosis of glaucoma remains a challenging task. Most individuals with glaucoma remain undiagnosed, while many healthy subjects are misdiagnosed and receive unnecessary treatment.

The lack of consensus on a reference standard for diagnosing glaucoma makes it difficult to evaluate newly proposed diagnostic tools. In recent years, advances in artificial intelligence (AI) have opened up the possibility for automated diagnoses of certain eye diseases using fundus photography. Deep learning (DL) algorithms have proven successful in detecting signs of diabetic retinopathy in fundus photos, achieving high accuracy when compared to a reference standard of human graders.[3] However, while the diagnosis of diabetic retinopathy is generally unequivocal, the same cannot be said for assessing the presence of glaucomatous damage, even for well-experienced glaucoma specialists.[4–6] Notably in the early stages of the disease, it can be very difficult, if not impossible, to ascertain the presence of glaucoma on a cross-sectional assessment of fundus photos. This is largely due to the wide variation in the appearance of optic discs in the normal population, as well as to the lack of a precise definition of what the “characteristic” features of glaucoma are. This lack of a clear definition of structural glaucoma questions the validity and usefulness of attempts to develop DL models to replicate human gradings for glaucoma diagnosis from fundus photos.

Spectral-domain optical coherence tomography (SDOCT) provides objective and reproducible quantitative assessment of neural loss in glaucoma. As an alternative to using human grading as the reference standard, DL models can be successfully trained to predict quantitative SDOCT measurements from analysis of fundus photographs. In a previous work, we developed a machine-to-machine DL algorithm that was able to provide continuous estimates of RNFL and neuroretinal rim thickness from fundus photos.[7, 8] The predicted measurements exhibited very high correlation to the observed SDOCT measurements, showing that the DL models could be used to automatically quantify neural loss from fundus photos.

Although SDOCT may be a reliable indicator of the presence of optic nerve damage, there are many other conditions that can lead to such damage, besides glaucoma. In addition, SDOCT by itself may be relatively unspecific for diagnosis of early glaucoma due to the wide variation in the normative measurement levels. The finding of correspondence between SDOCT and visual field abnormalities detected by standard automated perimetry (SAP) greatly enhances the specificity of diagnosis. However, to date, no objective definition of glaucomatous optic neuropathy (GON) has been proposed that incorporates both SDOCT and SAP.

In the present study, we propose an objective definition of GON using clearly defined structural and functional parameters from SDOCT and SAP. We then develop and validate a DL algorithm to detect GON on fundus photos, using the proposed objective definition as the reference standard. As such, to the best of our knowledge, the present work represents the first attempt to develop and validate an AI model to diagnose structurally and functionally defined GON using fundus photos.

## 2 Methods

This was a retrospective study that used cross-sectional data from the Duke Glaucoma Registry, a database of electronic medical and research records at the Vision, Imaging, and Performance Laboratory at Duke University. The Duke Health Institutional Review Board approved this study, and a waiver of informed consent was granted due to the retrospective nature of this work. All methods adhered to the tenets of the Declaration of Helsinki for research involving human subjects and the study was conducted in accordance with regulations of the Health Insurance Portability and Accountability Act.

The visual field tests were performed using SAP with the 24-2 Swedish Interactive Threshold Algorithm (Humphrey Field Analyzer II and III, Carl Zeiss Meditec, Inc., Dublin, CA) protocol. Unreliable tests with more than 33% fixation losses or more than 15% false-positive errors were excluded. RNFL thickness measurements were obtained from peripapillary circle scans, acquired using the Spectralis SDOCT (Software version 5.4.7.0, Heidelberg Engineering, GmbH, Dossenheim, Germany). According to manufacturer recommendations, tests with a quality score lower than 15 were excluded. The fundus photos present in the database were acquired from two different cameras: Nidek 3DX (Nidek, Japan) and Visupac (Carl Zeiss Meditec, Inc., Dublin, CA). The image was retained if the optic disc was entirely visible in the photo, and if no artifacts were present.

### Objective Definition of Glaucomatous Optic Neuropathy

GON was defined based on the presence of corresponding structural and functional damage on SDOCT peripapillary RNFL scans and SAP, respectively, acquired within six months of each other. The definition considered the possibility of both global as well as localized losses. An eye was considered to have GON if any of the following were present:

1. Global RNFL thickness outside normal limits and abnormal SAP as defined by Glaucoma Hemifield Test (GHT) outside normal limits or Pattern Standard Deviation (PSD) with P < 5%,
2. At least one sector in the superior RNFL thickness (temporal-superior or nasal-superior) outside normal limits with a corresponding abnormality on SAP inferior hemifield, defined as inferior hemifield mean deviation (MD) with P < 5%,
3. At least one sector in the inferior RNFL thickness (temporal-inferior or nasal-inferior) outside normal limits with a corresponding abnormality on SAP superior hemifield, defined as superior hemifield MD with P < 5%.

Superior and inferior SAP hemifield MD were calculated as the average of the total deviation values in each hemifield. The probability cutoffs for superior and inferior MD were derived from healthy subjects. Of note, as eyes with glaucoma may frequently have a dense arcuate defect occupying most of a hemifield, an approach using hemifield PSD to objectively characterize hemifield VF damage would have poor accuracy in diagnosis.

To be considered normal, or without GON, the SDOCT-SAP pair had to meet all of the following criteria:

1. Global RNFL thickness within normal limits,
2. RNFL thickness within normal limits for all sectors,
3. SAP GHT within normal limits,
4. SAP PSD probability “not significant”.

SDOCT-SAP pairs that did not meet criteria for GON or normal were considered suspects. These included eyes with only SDOCT abnormality or with only SAP abnormality. Although some suspects may in fact have early glaucoma, the lack of corresponding structural and functional damage leads to low specificity in the definition, which is undesirable in the context of developing an AI application for diagnosis and screening for glaucoma. Table 1 summarizes the criteria for the objective definition of GON.

**Table 1:** Summary of criteria for the objective definition of glaucomatous optic neuropathy (GON). Structural and functional criteria are derived from spectral-domain optical coherence tomography (SDOCT) and standard automated perimetry (SAP) results, respectively. To be considered glaucoma, it was necessary to meet the criteria for global or localized loss. To be considered normal, it was required that both SDOCT and SAP results were normal. SDOCT-SAP pairs that did not meet the criteria for either groups were considered suspects. *GHT = glaucoma hemifield testing; PSD = pattern standard deviation; MD = mean deviation*.

| Glaucomatous Optic Neuropathy |  |  |
| --- | --- | --- |
|  | SDOCT | SAP |
| **Global loss** | Global RNFL thickness outside normal limits | GHT outside normal limits or PSD $P < 5\%$ |
| **Localized loss** | RNFL thickness outside normal limits in at least one superior sector (temporal superior and/or nasal superior) | Inferior MD $P < 5\%$ |
| | RNFL thickness outside normal limits in at least one inferior sector (temporal inferior and/or nasal inferior) | Superior MD $P < 5\%$ |
| **Normal** |  |  |
| **Normal** | RNFL thickness within normal limits for all sectors and global | PSD probability not significant ($P > 5\%$) and GHT within normal limits |

### Deep Learning Model

A DL model was trained to predict the probability of GON on fundus photos, using as reference standard the proposed classification of GON defined based on SDOCT-SAP pairs. For each eye of each individual, all available fundus photos were matched with the closest SDOCT-SAP pair acquired within an interval of six months. Each SDOCT-SAP pair was classified as glaucoma, suspect or normal according to the definition above. Eyes that were suspected of glaucoma were not used for training the DL model.

The dataset was randomly divided at the subject level into 80% for training and validation of the model and 20% for final testing. The fundus photos were preprocessed, downsampled to 256 x 256 pixels and normalized so that all pixel values ranged between zero and one. Data augmentation, including horizontal flips, angle rotations, zoom and lighting changes of the original image, was performed to reduce the chance of overfitting and to improve the generalization of the algorithm.

Training of the DL algorithm was performed with transfer learning, using the Deep Residual Learning (ResNet50)[9] architecture, pre-trained with the ImageNet dataset. To adapt the architecture to our specific task, the last layer was replaced with a layer with output of size two: yielding probabilities of GON and normal. Initially, training was performed in the top layer, with the weights of the remaining layers frozen. Subsequently, all layers were unfrozen, and additional training was performed with differential learning rates, where smaller learning rates were used for earlier layers of the algorithm and larger rates for latter layers. The network was trained with minibatches of size 32, optimized with Adam.[10] The best learning rate was found using the cyclical learning method[11] with stochastic gradient descent with restarts.

By overlaying the Gradient-weighted class activation maps[12] with the input image, we built heatmaps that highlight the regions of the image that were most important for classification. This technique helps to visualize and understand the DL network predictions.

### Statistical analysis

The performance of the DL algorithm to detect GON on fundus photos was assessed in the test sample. Receiver operating characteristic (ROC) curves and the area under the ROC curve (AUC) were used to summarize the diagnostic accuracy. Furthermore, using a previously described ROC regression model,[13–15] ROC analysis was adjusted for age. Sensitivity and specificity were defined as the true positive and true negative rates, respectively. Due to the presence of multiple tests for each eye and each subject, comparison between groups was performed using a random effects mixed models,[16] and confidence intervals were derived with a bootstrap resampling procedure with resampling performed at the eye level.[17]

In order to investigate the performance of the DL algorithm in different levels of disease severity, SDOCT-SAP pairs were grouped into early, moderate and severe damage according to the Hodapp-Parrish-Anderson (HPA) classification of the SAP.[18] ROC curves, AUCs, sensitivity, and specificity were calculated for each level of disease severity.

Finally, interval likelihood ratios (LRs) were calculated to assess the impact of the results of the DL model in changing the probability of disease.[19] A LR is defined as the probability of a given test result in those with disease divided by the probability of that same test result in those without the disease.[20, 21] The application of LRs in the interpretation of results of imaging instruments for glaucoma diagnosis has been detailed previously.[22–24] LRs represent the best way to incorporate diagnostic test results in clinical practice according to the principles of evidence-based medicine. The LR for a given test result indicates how much that result will change the probability of disease, going from a pre-test probability (i.e., probability of disease before the test) to a post-test probability. A value of one means that the test provides no additional information, and ratios above or below one increase or decrease the likelihood of disease, respectively. Based on a prior classification definition, LRs greater than 10 or lower than 0.1 would be associated with large effects on post-test probability, LRs from 5 to 10 or from 0.1 to 0.2 would be associated with moderate effects, LRs from 2 to 5 or from 0.2 to 0.5 would be associated with small effects, and LRs closer to one would be insignificant.[21]

Development of the DL algorithm was performed in Python, while all statistical analyses were performed using Stata (version 15, StataCorp LP, College Station, TX). The alpha level (type I error) was set at 0.05.

## 3 Results

The dataset comprised of 9,830 fundus photos from 2,927 eyes of 2,025 individuals. The training/validation sample had 7,712 fundus photos from 2,342 eyes of 1,620 subjects, while the test sample had 2,118 fundus photos from 585 eyes of 405 subjects. Of the 585 eyes in the test sample, 305 (52%) had GON and 280 (48%) were normal. The median SAP MD were −7.5 and 0.24 dB for the glaucoma and normal groups, respectively (P = 0.000, random effects mixed model); while mean global RNFL thickness values were 67.0 and 98.3 *µ*m, respectively (P = 0.000, random effects mixed model). Table 2 shows demographic and clinical characteristics for the eyes in the study.

**Table 2:**
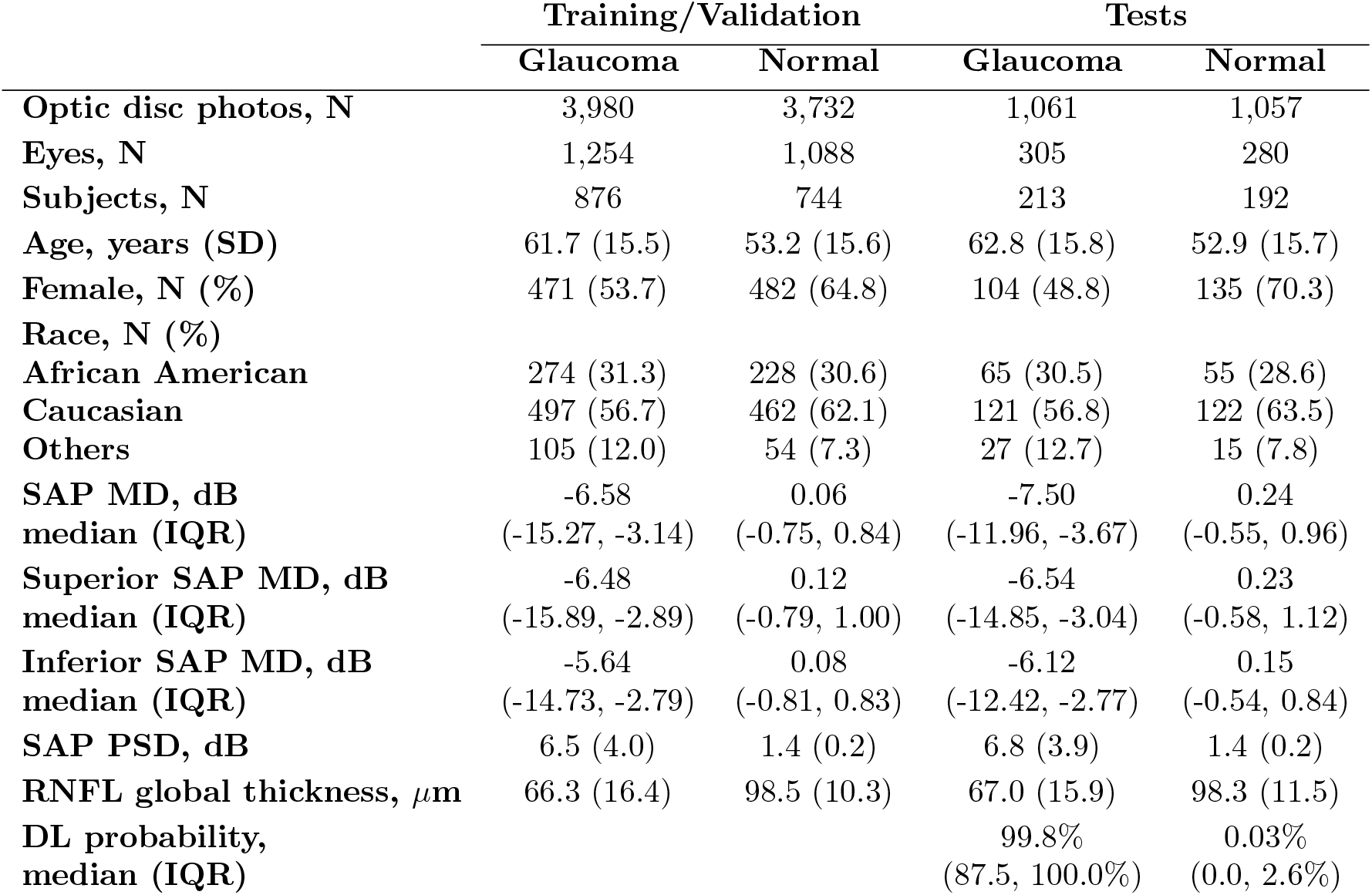
Demographic and clinical information of eyes and individuals included in the study. *SD = standard deviation; SAP = standard automated perimetry; MD = mean deviation; IQR = interquartile range; PSD = pattern standard deviation; RNFL = retinal nerve fiber layer; DL = deep learning*.

|  | Training/Validation |  | Tests |  |
| --- | --- | --- | --- | --- |
|  | Glaucoma | Normal | Glaucoma | Normal |
| **Optic disc photos, N** | 3,980 | 3,732 | 1,061 | 1,057 |
| **Eyes, N** | 1,254 | 1,088 | 305 | 280 |
| **Subjects, N** | 876 | 744 | 213 | 192 |
| **Age, years (SD)** | 61.7 (15.5) | 53.2 (15.6) | 62.8 (15.8) | 52.9 (15.7) |
| **Female, N (%)** | 471 (53.7) | 482 (64.8) | 104 (48.8) | 135 (70.3) |
| **Race, N (%)** |  |  |  |  |
| **African American** | 274 (31.3) | 228 (30.6) | 65 (30.5) | 55 (28.6) |
| **Caucasian** | 497 (56.7) | 462 (62.1) | 121 (56.8) | 122 (63.5) |
| **Others** | 105 (12.0) | 54 (7.3) | 27 (12.7) | 15 (7.8) |
| **SAP MD, dB** | -6.58 | 0.06 | -7.50 | 0.24 |
| **median (IQR)** | (-15.27, -3.14) | (-0.75, 0.84) | (-11.96, -3.67) | (-0.55, 0.96) |
| **Superior SAP MD, dB** | -6.48 | 0.12 | -6.54 | 0.23 |
| **median (IQR)** | (-15.89, -2.89) | (-0.79, 1.00) | (-14.85, -3.04) | (-0.58, 1.12) |
| **Inferior SAP MD, dB** | -5.64 | 0.08 | -6.12 | 0.15 |
| **median (IQR)** | (-14.73, -2.79) | (-0.81, 0.83) | (-12.42, -2.77) | (-0.54, 0.84) |
| **SAP PSD, dB** | 6.5 (4.0) | 1.4 (0.2) | 6.8 (3.9) | 1.4 (0.2) |
| **RNFL global thickness, $\mu\text{m}$** | 66.3 (16.4) | 98.5 (10.3) | 67.0 (15.9) | 98.3 (11.5) |
| **DL probability,** |  |  | 99.8% | 0.03% |
| **median (IQR)** |  |  | (87.5, 100.0%) | (0.0, 2.6%) |

The median DL probability of glaucoma (interquartile range [IQR]) assigned to fundus photos with GON and normal were 99.8% (87.5, 100.0%) and 0.03% (0.0, 2.6%), respectively (P = 0.000, random effects mixed model). Figure 1 illustrates the distribution of DL probabilities. The DL algorithm had an age-adjusted AUC of 0.92 (95% CI: 0.88, 0.95) in the test sample. For a 95% specificity, the model had 77.3% sensitivity.

**Figure 1:**
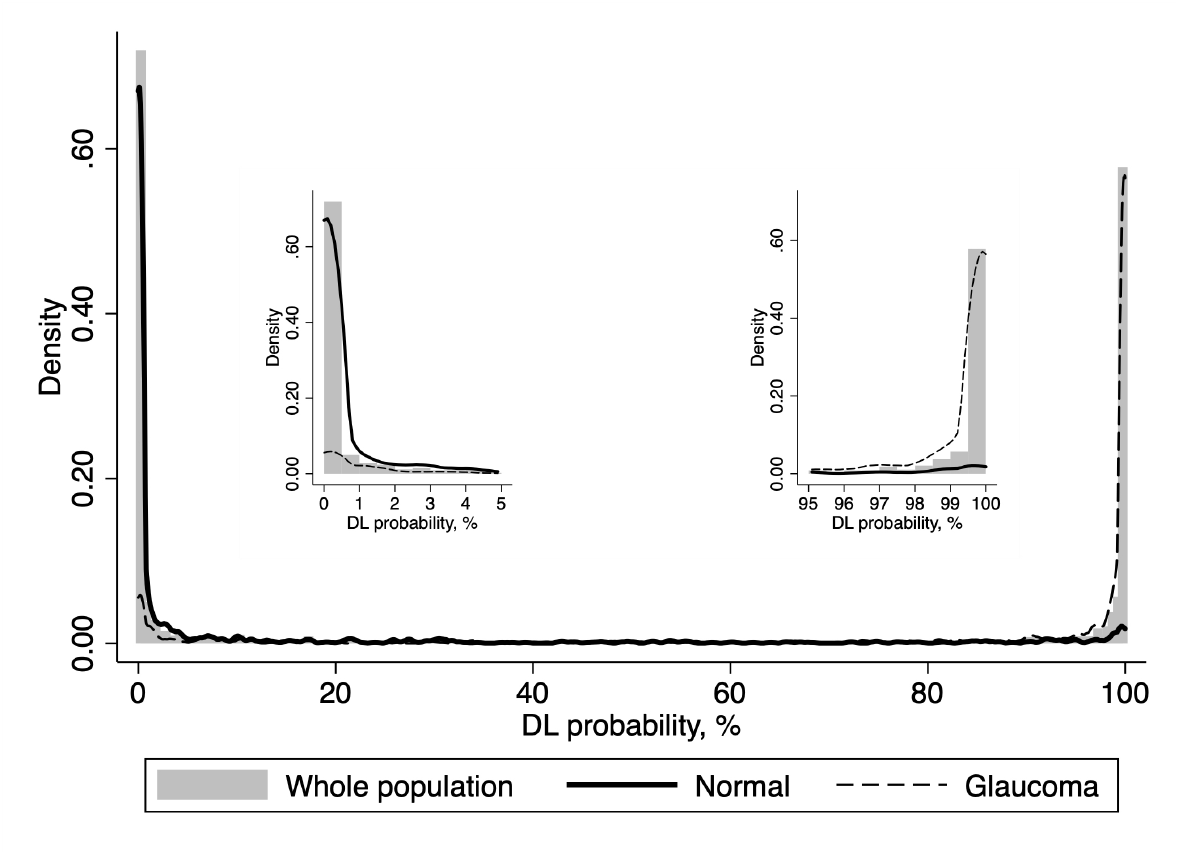
Distribution of the deep learning (DL) probabilities of glaucoma for the photos included in the test sample. The overall distribution is represented by shaded bars. Tails of the distribution (left, low probability of glaucoma; and right, high probability of glaucoma by the DL algorithm) are shown in detail. Photos classified as normal by the objective criteria proposed in the study had DL probabilities of glaucoma concentrated closer to zero (solid line). For photos classified as glaucoma, the DL probabilities were concentrated closer to one hundred (dashed line).

The AUC values increased with worse levels of disease severity, achieving maximal age-adjusted AUC of 0.96 and sensitivity of 85.1% (at 95% specificity) among eyes with severe glaucoma. In Figure 2 and Table 3, ROC curves and AUC values, as well as sensitivities at 95% specificity, are presented across levels of disease severity. Figures 3 and 4 illustrate eyes included in the study, classified as normal and as glaucomatous, respectively.

**Table 3:**
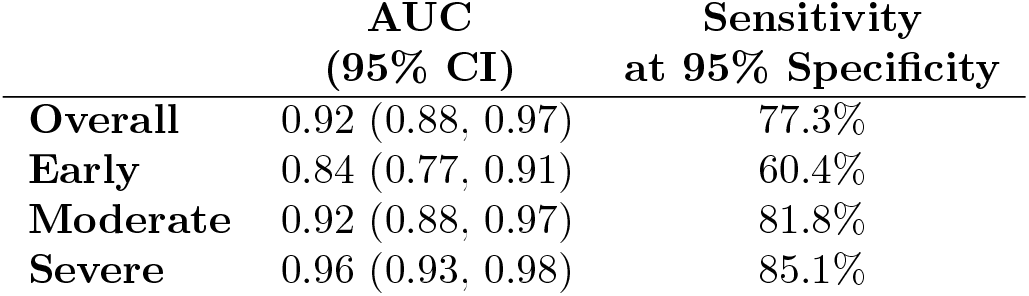
Area under the receiver operating characteristic curve (AUC) and sensitivity at 95% specificity for different levels of disease severity, according to the Hodapp-Parrish-Anderson classification of glaucoma severity, based on visual field damage. *CI = confidence interval*

**Figure 2:**
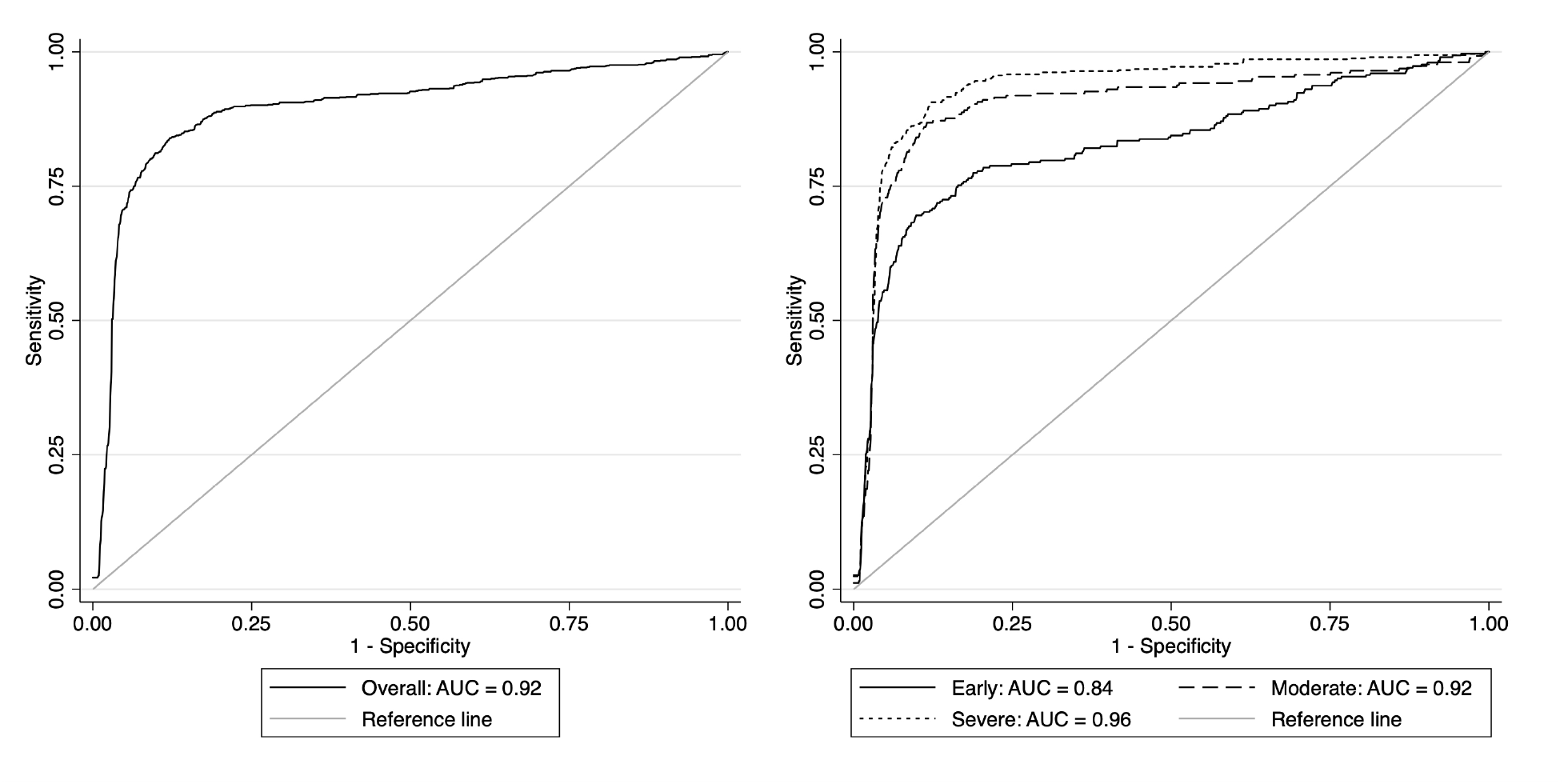
Age-adjusted receiver operating characteristic (ROC) curves illustrating the ability of the algorithm to discriminate between normal and glaucoma for all photos (left) and at different levels of disease severity (right), according to the Hodapp-Parrish-Anderson classification of severity, based on visual field damage. *AUC = area under the ROC curve*

**Figure 3:**
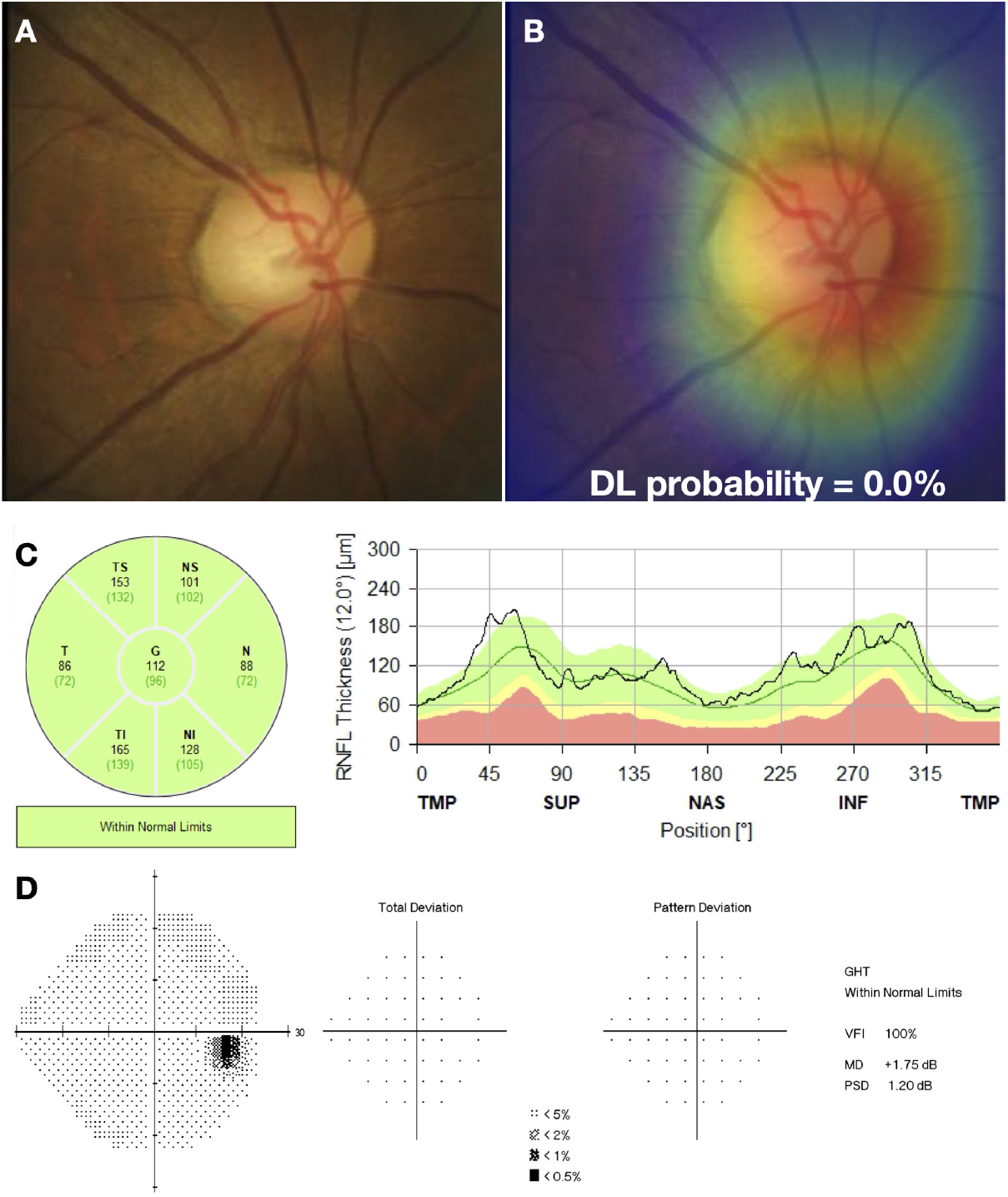
Example of a normal case included in the study. (A) illustrates the fundus photo used as input for the deep learning (DL) algorithm, which predicted a glaucoma probability of 0.0%. (B) shows the regions that were most important for the classification, in which the heatmap diffusely highlights the optic disc. (C) illustrates the spectral domain optical coherence tomography and (D) the standard automated perimetry, both without abnormalities.

**Figure 4:**
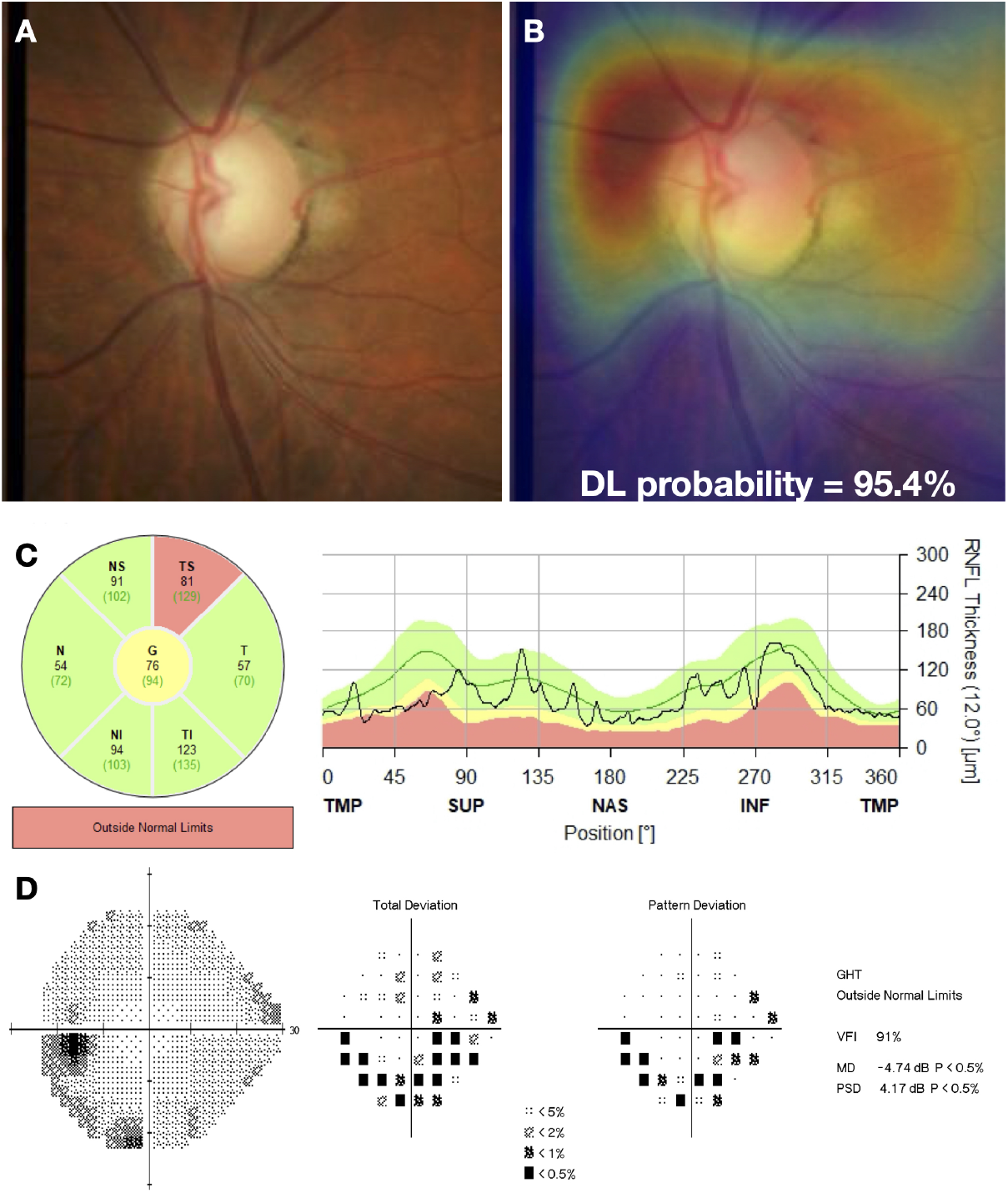
Example of a glaucoma case included in the study. (A) illustrates the fundus photo used as input for the deep learning (DL) algorithm, which predicted a glaucoma probability of 95.4%. (B) shows the regions that were most important for the classification, in which the heatmap highlights the superior half of the optic disc and peripapillary region. (C) illustrates the spectral domain optical coherence tomography with a temporal superior retinal nerve fiber layer defect, with a corresponding inferior arcuate defect in the standard automated perimetry (D).

The LRs are presented in Table 4 and help identify meaningful intervals of the DL probabilities for the risk of the disease. Test results with DL probability *>* 99% or *>* 99.9% were associated with large changes in increasing the probability of disease (LRs of 10.4 and 38.2, respectively); whereas test results with DL probability < 0.1% were associated with large change in decreasing the probability of disease (LR of 0.06). Any of these test results would provide strong evidence regarding the presence or absence of GON. As Table 4 shows, other test results would also increase or decrease the probability of disease, but provide less conclusive evidence, which could still be helpful depending on the setting of application of the test.

**Table 4:** Likelihood ratios for different intervals of deep learning (DL) probabilities of glaucoma.

| DL probability<br>% | Glaucoma<br>N (%) | Normal<br>N (%) | Total<br>N (%) | Likelihood<br>Ratio |
| --- | --- | --- | --- | --- |
| 0 – 0.1 | 38 (3.6) | 604 (57.1) | 642 (30.3) | 0.06 |
| 0.1 – 1 | 32 (3.0) | 141 (13.3) | 173 (8.2) | 0.23 |
| 1 – 10 | 57 (5.4) | 116 (11.0) | 173 (8.2) | 0.49 |
| 10 – 50 | 70 (6.6) | 86 (8.1) | 156 (7.4) | 0.81 |
| 50 – 90 | 78 (7.4) | 47 (4.4) | 125 (5.9) | 1.65 |
| 90 – 99 | 141 (13.3) | 36 (3.4) | 177 (8.4) | 3.90 |
| 99 – 99.9 | 146 (13.8) | 14 (1.3) | 160 (7.6) | 10.4 |
| 99.9 – 100 | 499 (47.0) | 13 (1.2) | 512 (24.2) | 38.2 |

A total of 4,899 fundus photos from 1,648 eyes of 1,085 individuals were classified as suspect. When applied to this group, the DL algorithm predicted a median probability of glaucoma (IQR) of 6.4% (0.2, 82.4%). These eyes had median (IQR) SAP MD of −1.82 dB (−4.32, −0.31) and mean (SD) global RNFL thickness of 94.4 *µ*m (15.7). When suspect eyes were divided in quartiles according to the DL probability, those in the highest quartile (higher probability) had significantly lower global RNFL thickness measurements than those in the lowest quartile [90.3 *±* 16.8 vs. 98.7 *±* 14.2 *µ*m; P = 0.019] and also worse SAP MD, although the difference was not statistically significant −2.3 (−5.4, −0.7) vs. −1.2 (−3.3, 0.2); P = 0.390]. Supplementary Figures S1 and S2 illustrate eyes that were classified as suspects and their corresponding predictions by the DL algorithm.

## 4 Discussion

In the present study, we developed novel objective criteria for defining GON, based on the correspondence of structural and functional disease measures. We then used the proposed reference standard to train a DL algorithm to estimate the probability of GON from optic disc photos. The algorithm was able to accurately discriminate photos from eyes with and without GON based on the proposed reference standard, suggesting that DL analysis of fundus photos may have a role in screening for glaucomatous damage. Additionally, our results provide support for the use of an objectively defined structural and functional reference standard for GON, given the strong agreement found between DL predictions and the reference classifications.

Fundus photos are an inexpensive and portable method to image the optic disc. With recent advances in AI, automated algorithms have been developed to analyze fundus photos and yield a probability of GON. However, in most previous works, the development of DL algorithms relied on human gradings to establish the reference standard, an approach that presents severe limitations. First, human gradings tend to over- and underestimate the likelihood of glaucoma,[25] and, even among specialists, there is poor reproducibility and low interobserver agreement.[4–6] Second, labelling a large number of fundus photos to be used as reference standard is cumbersome and time consuming. Third, the experience of the evaluators and the criteria used to define GON varied tremendously across those studies. Finally, the accuracy achieved using the reference standard based on human gradings does not necessarily translates well to clinical practice. As an example, in a work by Phene et al.,[26] a DL algorithm developed to detect GON, defined by human gradings, in fundus photos, achieved an AUC of 0.945. When evaluated in samples with a different reference standard, defined by a complete glaucoma workup and by International Classification of Diseases codes, the AUCs were 0.855 and 0.881, respectively.

In order to avoid subjective biases and low accuracy from human labelling, we developed an objective definition of GON, based on quantitative metrics of both SAP and SDOCT. SDOCT measurements of RNFL thickness are objective, reproducible, and are considered the standard of care for quantification of structural damage in glaucoma.[27] SAP is used to detect and classify levels of severity of functional loss18 and is correlated to quality of life and visual disability.[28] Since both tests are metrics of the same disease, using a combination of their results increases the specificity of the classification – which is particularly important for a reference standard used in developing algorithms aimed at screening for glaucoma, a disease that has a relatively low-prevalence. To the best of our knowledge, our study is the first to propose such objective definition of GON based on SDOCT and SAP.

The DL algorithm predicted probabilities of GON with high confidence, with a median DL probability of 0.03% (IQR: 0.0, 2.6%) for photos classified as normal and 99.8% (IQR: 87.5, 100.0%) for photos classified as glaucomatous. These results not only provide confidence on the usefulness of the DL classifier, but also provide internal validation for our proposed objective definition of GON. When applied to photos of eyes that had been classified as suspects, the median DL probability was 6.4% (IQR: 0.2, 82.4%), with a wide distribution of predictions. This is likely due to the fact that some eyes with pre-perimetric glaucoma, as well as some normal eyes, were included as suspects given the lack of correspondent structural and functional damage. In fact, when we compared structural and functional measurements in glaucoma suspect eyes divided by quartiles of DL probability, we found that those in the highest quartile had significantly lower global RNFL thickness measurements suggesting that the model was capturing those suspects at higher risk. The age-adjusted AUC of the DL probabilities was 0.92, with a 77.3% sensitivity at 95% specificity.

When evaluated in different levels of disease severity, the DL algorithm performance improved with worse severity (Early: AUC = 0.84, Moderate: AUC = 0.92, Severe: AUC = 0.96), correctly identifying more than 60% of the early cases and over 80% of the moderate and severe cases at a specificity of 95%. This performance makes the DL algorithm particularly useful for screening of moderate and severe glaucoma, which require prompt referral, but also with a high specificity, so as not to overwhelm the healthcare system with false-positive referrals. One could argue that the algorithm still misses about 20% of moderate and severe cases of glaucoma. However, given that previous population-based studies show that over 90% of patients with glaucoma remain undiagnosed in developing areas,[29, 30] such a simple and inexpensive approach could potentially help reduce the burden of disability from glaucoma.

We also presented LRs for different intervals of DL probabilities. In contrast to sensitivity and specificity, which are difficult to apply in practice, LRs can be readily incorporated into clinical decision-making. In our work, DL probability values lower than 0.1% or greater than 99% were associated with large changes in the post-test probability, providing major conclusive changes regarding presence or absence of damage from glaucoma. To illustrate, consider the application of the test in a situation of screening, where the overall prevalence of glaucoma would be estimated at 5%.[29, 30] This would then be considered the pre-test probability. If this patient is tested and gets a result with DL probability greater than 99.9% (LR of 38.2), this would bring the post-test probability to 66%, indicative of the need for referral.

The calculation of LRs helps to understand the usefulness of the test under different circumstances as well. For example, when applied to the evaluation of suspects in clinical practice or during opportunistic screening, levels of pre-test probability of disease would often be significantly higher, as those patients would most likely have clinical findings raising the probability of disease, such as family history or suspicious optic disc appearance. For example, if a patient has a pre-test probability of 50%, a test result with DL probability of 0.1% (LR = 0.06) would bring the post-test probability to just 5.7%. On the other hand, if a test result with DL probability of 99% is obtained, the post-test probability would jump to 91%. DL probabilities below 0.1% or higher than 99% occurred in 62% of the overall sample, indicating that the test would provide strong evidence in a majority of patients.

This study has limitations. Our cohort consisted of a clinic-based population, meaning that further validation in real-world settings, for population-based or opportunistic screening, is necessary. Also, the classification did not include risk factors, such as intraocular pressure, race, family history, and age. However, such information can be aggregated to estimate the pre-test probability and then one can apply the DL model to modify such probability. Finally, the DL algorithm would benefit from additional external validation on a population from a different geographical area.

In conclusion, we developed a DL algorithm to evaluate fundus photos for glaucomatous damage, based on a novel reference standard definition of GON that uses both structural and functional measures of disease. We demonstrated that the DL algorithm can produce large changes in the probability of disease in the majority of subjects, supporting its usefulness for decision-making. Furthermore, our proposed objective reference standard to define GON may increase the comparability of diagnostic studies across devices and populations, helping to improve the development and assessment of diagnostic tests in clinical practice.

## Data Availability

The authors confirm that the data supporting the findings of this study are presented within the article and its supplementary materials

## Funding supports

Supported in part by National Institutes of Health/National Eye Institute grant EY029885.

## Author contributions

EBM, AAJ and FAM conceived and designed the study described here. EBM, SIB and FAM performed statistical analyses. EBM and AAJ collected and managed the data. IMT and FAM advised in study design, data analysis methods and drafting the manuscript. All authors reviewed and revised the manuscript prior to submission.

## 5 Supplementary Material

**Figure S1:**
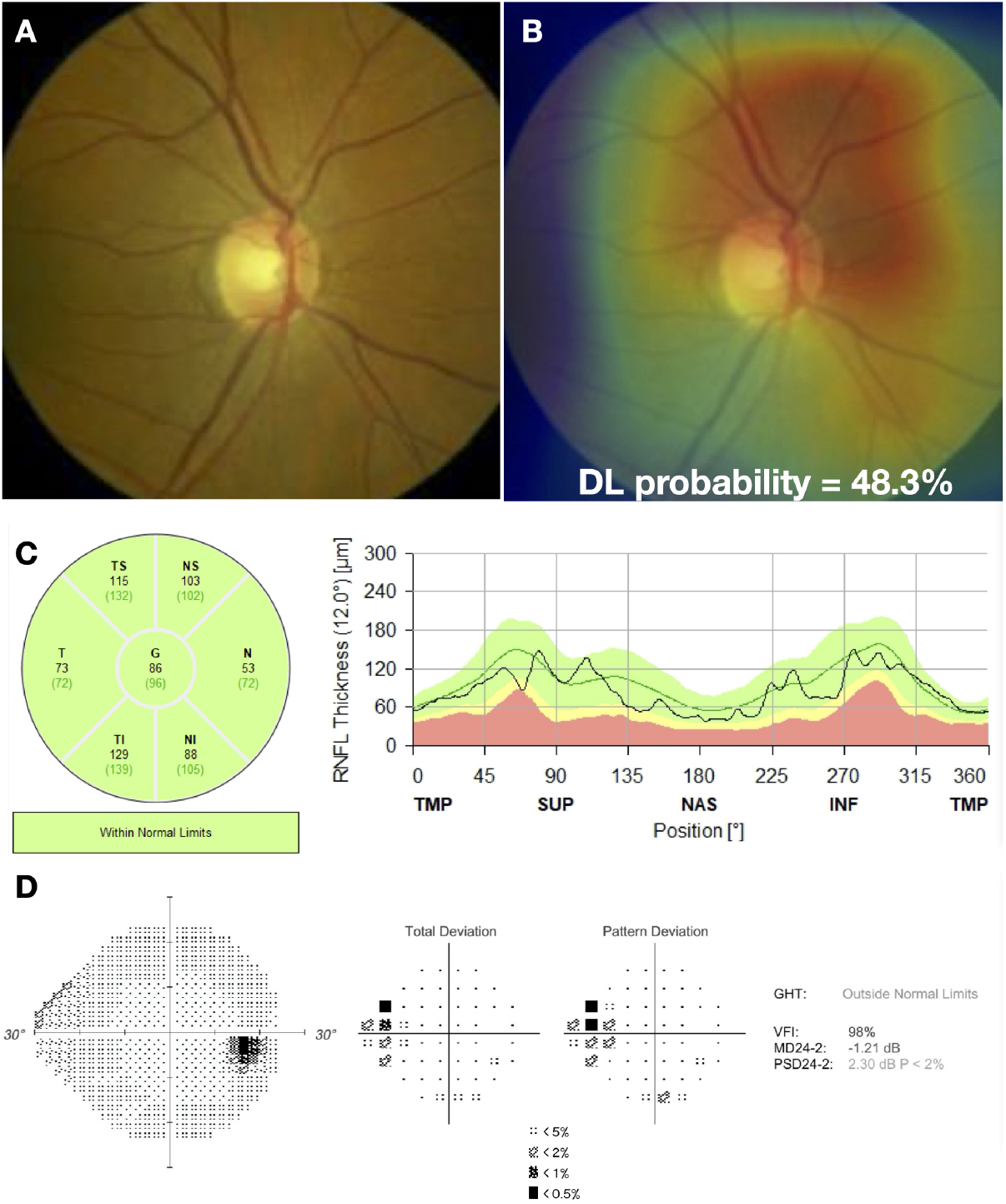
Example of a case classified as suspect. (A) illustrates the fundus photo used as input for the deep learning (DL) algorithm, which predicted a glaucoma probability of 48.3%. (B) shows the regions that were most important for the classification, in which the heatmap highlights diffusely the image, but mostly the superior half of the optic disc and the peripapillary region. Although the global and sectoral retinal nerve fiber layer thickness are within normal limits in the spectral domain optical coherence tomography (C), there is an apparent superior nasal step defect and the pattern standard deviation (PSD) is abnormal (P ¡ 5%) in the standard automated perimetry (D).

**Figure S2:**
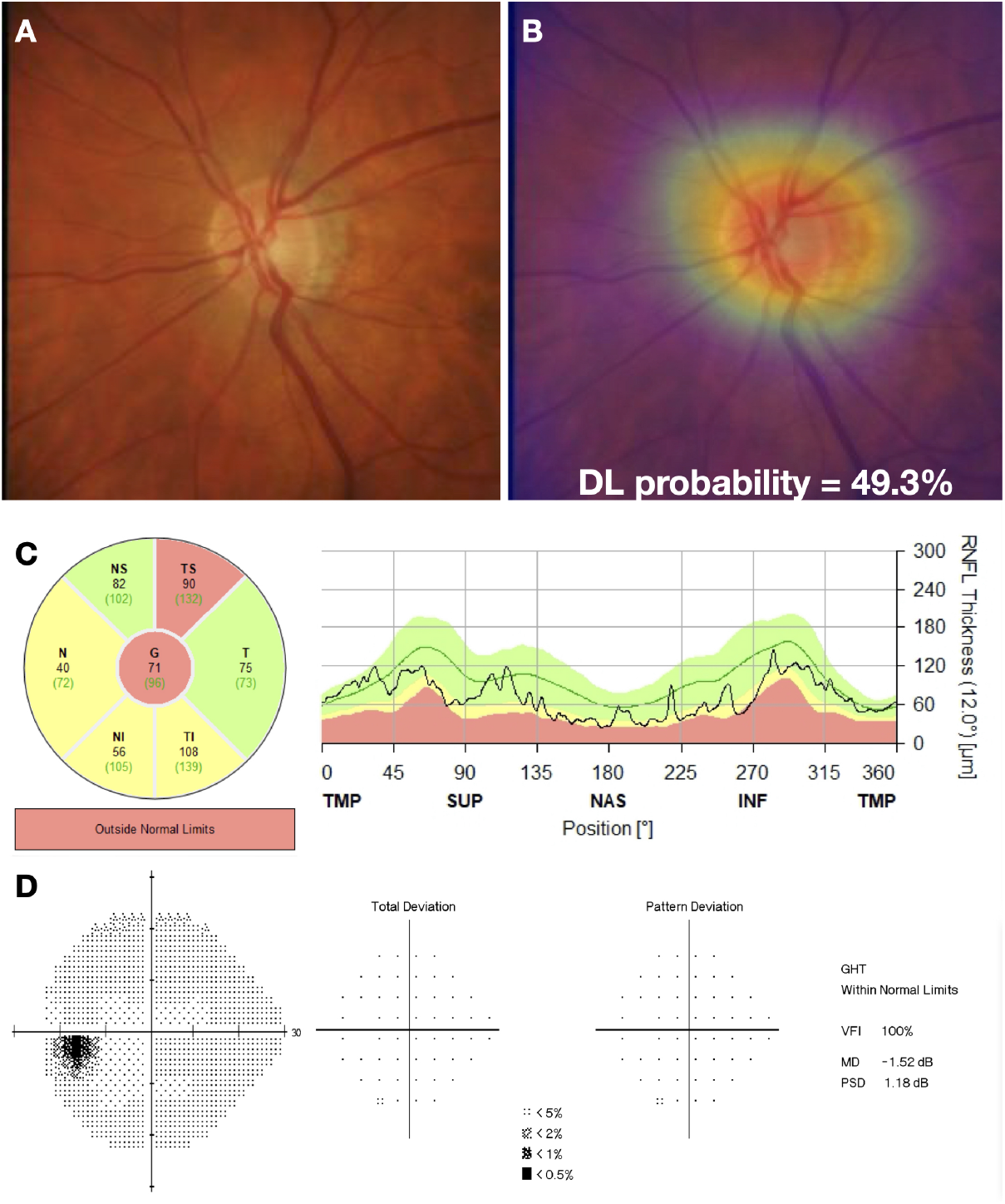
Example of a case classified as suspect. (A) illustrates the fundus photo used as input for the deep learning (DL) algorithm, which predicted a glaucoma probability of 49.3%. (B) shows the regions that were most important for the classification, in which the heatmap highlights diffusely the optic disc and a peripapillary atrophy. Although the spectral domain optical coherence tomography (C) is abnormal (global and temporal superior sectors outside normal limits, and 3 other sectors borderline), there is no defect in the standard automated perimetry (D).

